# Peer support boosted Hepatitis C treatment access among marginalised populations in England: A Bayesian causal factor analysis

**DOI:** 10.64898/2026.04.20.26351261

**Authors:** Constantin Schmidt, Pantelis Samartsidis, Shaun R. Seaman, Beatrice Emmanouil, Graham R. Foster, Leila Reid, Stuart Smith, Daniela De Angelis

## Abstract

To minimise health disparities, equitable access to medical treatment is paramount. In a pioneering intervention, National Health Service England’s Hepatitis C virus (HCV) programme has implemented country-wide peer support to boost treatment access. Peer support workers (*peers*) are individuals with relevant lived experience, who promote testing and treatment in marginalised populations underserved by traditional health services. We evaluated the English peers intervention, exploiting its staggered rollout and rich surveillance data between June 2016 and May 2021. Peers increased HCV cases identified by 13·9% (95% credible interval (95% CrI) [5·3, 21·7]), sustained viral responses by 8·0% (95% CrI [–4·4, 18·6]), and drug services referrals by 8·8% (95% CrI [–12·5, 22·6]). The intervention’s effectiveness was magnified during the first COVID-19 lockdown and individuals supported by peers typically belonged to populations with poor treatment access. Our findings indicate that peers can boost equity in treatment access on a national scale.

## 1 Introduction

Hepatitis C virus (HCV) is a blood-borne virus affecting the liver. Chronic HCV infection can lead to acute liver damage, cirrhosis, cancer, and eventually death. ^1^ HCV infections are a major global health concern. In 2022, it was estimated that approximately fifty million people globally were living with chronic HCV infection. ^2^

HCV disproportionally affects marginalised populations, which tend to be excluded from traditional health services. The most notable example is people who inject drugs (PWID). ^3^ In 2022, approximately 5·8 million PWID, about 38·8% of the global population of PWID, were infected with HCV. ^4^ In high-income countries, other factors associated with heightened risk of HCV infection include a history of incarceration, experiencing homelessness, or having grown up in a country with high HCV infection prevalence. ^1^

Global efforts to reduce the burden of HCV are ongoing. Countries committed to the World Health Organization’s (WHO) 2030 hepatitis elimination targets aim to reduce HCV infection incidence by 80% and HCV-related mortality by 65% by 2030 compared to 2015 levels. ^5^ Paramount to achieving the WHO’s HCV elimination targets is the widespread and equitable administration of direct-acting antiviral (DAA) HCV treatments. ^6^ Since 2015, DAAs have become the standard of care, achieving sustained viral response in 95% of treated patients after an 8–12-week oral course with few adverse effects. ^7^

The United Kingdom has emerged as one of the global pioneers in HCV elimination. It was one of seven countries in which the WHO’s HCV strategy was piloted. ^5^ Treatment with DAAs is provided free of charge through the National Health Service (NHS). In England, treatment provision is handled regionally by 22 *Operation Delivery Networks* (ODNs). Various interventions have been introduced to engage marginalised populations to HCV treatment, including treatment as prevention, testing and treatment by pharmacists, and testing and treatment in mobile units placed in known homeless population areas. ^8–10^ Still, the burden of HCV remains significant in England where, in 2023, an estimated 55,900 adults were living with a chronic HCV infection, 84·6% of whom were PWIDs or people with a history of injecting drug use. ^1,11^

To ensure further progress towards HCV elimination, NHS England (NHSE) partnered with the pharmaceutical industry to set up a country-wide peer support programme. Peer support workers (*peers*) are individuals from the community with relevant lived experience of HCV, who raise awareness of the virus, encourage testing and treatment, and help navigate the treatment process. Starting in 2018, peers were trained by the *Hepatitis C Trust* (HCT), a London-based charity. In order to not overwhelm the HCT’s capacity, peers were introduced in staggered fashion across all ODNs. The NSHE peers intervention was pioneering in its scale and integration with local treatment providers as every individual treated by NHSE for HCV was eligible to receive peer support.

Peer support has emerged as a widely used intervention to improve treatment uptake, particularly in disease areas that disproportionately affect marginalised populations. In the context of Human Immunodeficiency Virus (HIV), a recent meta-analysis of 10 randomised controlled trials (RCTs) found that peer support increased the uptake of HIV testing services. ^12^ Peers seemed to also be effective in outreach campaigns via social media. ^13^ In the context of tuberculosis (TB) the evidence for peer support is mixed. A systematic review found that direct incentives were more effective than peers in promoting TB screening in homeless populations. ^14^ In addition, peers did not increase the uptake of TB screening by a mobile X-ray unit in homeless hostels in London compared to outreach done by the mobile unit’s staff. ^15^ Evidence on peer support in the context of HIV and TB remains confined to targeted small-scale initiatives.

Evidence on peer support in the context of HCV remains sparse (see search strategy in Appendix A). Qualitative studies conducted in the UK, ^16,17^ Georgia, ^18^ and Australia ^19^ suggest that peer support may be a valuable intervention to facilitate HCV treatment uptake. Results from two RCTs indicate that peer support may increase the probability that individuals who test positive for HCV infection start treatment. ^20,21^ However, these RCTs were relatively small. One observational study from England found that peer support increased the number of HCV treatment initiations and the odds of treatment completion. ^22^ However, this study covers only the first two months after peer introduction, limiting conclusions about longer-term effects. In addition, no study provides evidence on whether peers are able to engage with the marginalised populations most at risk of an HCV infection.

Our study fills a critical gap in the literature by being the first to evaluate the long-term effects of a country-wide peer support intervention aimed at improving treatment related outcomes in marginalised populations. Our primary outcome of interest was the number of HCV-infected individuals identified (*HCV cases identified*), as no previous study had considered this outcome. Our secondary outcomes were the number of individuals completing DAA treatment and achieving confirmed sustained viral response (*SVR*) and the number of individuals initiating DAA treatment that are referred from drug services (*drug service referrals*, see Table 4 in Appendix). We also investigated how the effects of the peers intervention changed with the number of peer-months of exposure since intervention start and during the first national lockdown in England, a period when health inequalities were exacerbated. In addition, our results provide insights into the demographics of individuals engaging with peers. Our study was enabled by high-quality HCV surveillance data from the NHS and HCT. Exploiting the staggered adoption of the peers intervention as a natural quasi-experiment, we employed a state-of-the-art causal inference method, which mitigates the risk of confounding bias common to observational studies. ^23^

## 2 Methods

### Intervention implementation

Full details regarding the implementation of the peer programme have been given elsewhere, ^22^ we provide a brief overview. The peers programme funding was established through a partnership between NHSE and the pharmaceutical industry. Beginning in January 2018, peers were trained by HCT, the United Kingdom’s only charity dedicated to HCV. The majority of HCT’s staff and volunteers have relevant lived experience, including experience of HCV, homelessness, injecting drug use and/or the criminal justice system. HCT staff also have professional experience and skills in engagement, outreach and care.

To implement national coverage and prevent ovewhelming HCT capacity a staggered approach to implementation was taken whereby 4 ODNs were provided the peers service on a quarterly basis. The order of implementation times was also determined by each ODN’s administrative capacity and willingness to introduce a peer. Later, many ODNs decided to employ additional peers made available by further NHSE funding.

Peers operate in partnership with services supporting populations more likely to contract HCV such as drug services, homeless hostels and community criminal justice settings. In these settings, peers provide educational workshops and deliver HCV and other blood-borne virus testing. People diagnosed with HCV by peers or NHS teams are offered ongoing peer support. This support is tailored to individual needs and may include appointment reminders, accompaniment to appointments, medication delivery, and well-being support. Peers might also provide essential healthcare interventions such as needle exchange, safer injecting advice, and signposting to other services. In addition, peers recruited volunteers to boost outreach efforts.

### Outcomes, exposure, and data sources

We evaluated the effect of peers support on three outcomes: (i) the number of HCV-infected individuals identified (*HCV cases identified*); (ii) the number of individuals completing DDA treatment and achieving confirmed sustained viral response (*SVR*); and (iii) the number of individuals initiating DDA treatment that are referred from drug services (*drug service referrals*, see Table 4 in Appendix B for more detailed definitions). The number of HCV cases identified was our primary outcome, as no previous study had considered this outcome. An ODN was defined exposed to the intervention if at least one peer was employed, and the cumulative number of peer-months of exposure served as a measure of exposure intensity. Using these data, we constructed a longitudinal panel with monthly observations of all ODNs covering June 2016 to May 2021.

The number of HCV cases identified was calculated using *Blueteq high-cost drugs system* (Blueteq), a web-based platform used by NHSE for the management of high-cost prescription medicines. ^24^ Before providers order DAA medication, a treatment funding request needs to be approved through Blueteq. Without an approved funding request, providers will not be reimbursed. Since all HCV infected individuals are DDA treatment eligible, Blueteq is thus an exhaustive database of HCV infected individuals identified by NHSE. Individual information relevant to treatment — age, treatment history, cirrhotic status, and treatment provider — are recorded.

SVRs and drug service referrals were retrieved from the *Hepatitis C patient registry and treatment outcome system* (the Registry), a secure online system containing data on people treated for HCV infection in England. ^25^ The system is updated daily by staff in the ODNs. The Registry only contains a subset of the patients recorded in Blueteq, but records more information on each patient. It includes demographic details (age, gender, ethnicity, country of birth), liver disease stage, likely route of transmission, relevant comorbidities (HIV co-infection, history of liver transplants), treatment details (initiation date, completion data, treatment given) and source of referral (general practitioner, drug service, *etc*).

Information on the activities of the peers was retrieved from the secure HCT database. The database contains the exact date each peer started working and the ODN they worked in. Further, it contains characteristics of each HCV-infected individual supported by peers, including some demographic information (gender, age) and some risk factors for HCV infection (whether they are currently injecting, homeless, *etc*).

### Statistical analysis

To evaluate the peers intervention, we compared observed outcomes to intervention-free counterfactuals. Intervention-free counterfactuals are defined as the outcomes that would have been observed if the peers intervention had not been implemented. Thus, the causal effect of the intervention is the difference between the observed outcome and the counterfactual; we call this difference the individual treatment effect (IIE). We obtained an IIE for each combination of outcome, each exposed ODN, and each post-intervention time period. IIEs were then used to calculate four summary intervention effects of interest: the overall change in each outcome due to the intervention, defined as the sum of IIEs across all post-intervention periods; the overall percentage change in each outcome, defined as the overall change in the outcome divided by the sum of intervention-free counterfactuals; the change in each outcome during the COVID-19 lockdown, defined as the sum of IIEs across post-intervention periods during which the first English COVID-19 lockdown was in place; and the percentage change in each outcome during the COVID-19 lockdown, defined as the change in the outcome during the COVID-19 lockdown divided by the sum of intervention-free counterfactuals. As further causal effect of interest, we examined rate ratios (RR). For each outcome, RRs were defined as the ratio of the mean of the outcome for a given intervention intensity to the mean of the outcome under no intervention. We used RR to investigate the relationship between exposure intensity and intervention effect. We provide mathematical definitions of these intervention effects in AppendixD.

The intervention-free counterfactuals can never be observed for post-intervention time periods and thus needed to be imputed. The statistical model used to impute these counterfactuals needed to be flexible enough to model underlying temporal trends in the data and to adjust for confounding. In our study, confounding might be caused by the fact that peer start times were based on the willingness and capacity of an ODN to integrate peers, which might also affect the outcomes. To carry out the imputation we employed a recently developed Bayesian causal factor model (Appendix D). ^23^ Briefly, we assumed that the data arise from a Negative Binomial distribution, with mean modelled as the sum of four terms: (i) an ODN-specific intercept to control for underlying, time-constant differences between ODNs, such as willingness and capacity of an ODN to integrate peers; (ii) a time-fixed effect to adjust for temporal trends common to all ODNs which might, for example, be caused by general changes in HCV prevalence; ^26,27^ (iii) interactive fixed effects to account for the time-varying effect of a set of ODN-specific unmeasured confounders; (iv) a term capturing the impact of the peers intervention on the mean of the outcome, which depends on the number of peer-months an ODN has been exposed and on whether a COVID-19 lockdown is in place. We accounted for additional variability in outcomes due to the intervention by allowing the model parameter describing overdispersion to differ between periods with and without active peers. We note that our empirical strategy relaxes the parallel-trends assumption made by the widely employed linear difference-in-differences method.

The models were fitted under the Bayesian paradigm. This allowed us to obtain the posterior distributions of RRs and IIEs. IIEs were then aggregated into the summary intervention effects of interest. We used the mean of the posterior distribution of an intervention effect as the point estimate, and the 2·5th and 97·5th centiles as the bounds of a 95% posterior credible interval (95% CrI). We further calculated the probability that an intervention effect is greater than zero; this will be referred to as the posterior probability of a positive intervention effect (PPos). All analyses were conducted in the `R` library `rstan`. ^28^

## 3 Results

### Data descriptives

After cleaning, the final sample included 56,137 HCV-infected individuals in Blueteq and 44,637 HCV-infected individuals in the Registry (see Figures 4 and 5 in Appendix B for eligibility criteria). Of these, 15,574 (Blueteq) and 11,509 (Registry) were managed during a time period when peers were operating within their ODN. There was a substantial amount of missing values in the variables ethnicity (4,169 [9.3%] out of 44,637 individuals, Registry); drug use (9,943 [22.3%] out of 44,637 individuals, Registry); and treatment history (5,587 [10.0%] out of 56,137 individuals, Blueteq). The majority of individuals was non-cirrhotic (44,900 individuals [80.3% of non-missing cases] in Blueteq and 35,485 individuals [76·4% of non-missing cases] in the Registry; Table 1). In Blueteq, 43,717 individuals (86·5% of non-missing cases) were HCV treatment-naïve. In the Registry, 11,742 individuals (33·8% of non-missing cases) were current drug users and 13,869 (39·9% of non-missing cases) were past drug users. The majority of individuals recorded in the Registry were male (31,427 [70·6% of non-missing cases]), while white ethnicity was the most common (33,721 [83·5% of non-missing cases]).

**Table 1:** Characteristics of study sample. Results are shown as count (%). Characteristics are for HCV-infected individuals recorded in the Blueteq dataset (left) and the English national Hepatitis C treatment registry (right) between June 2016 and May 2021. Percentages were calculated using non-missing observations.

|  | **Blueteq** |  | **Registry** |  |
| --- | --- | --- | --- | --- |
|  | *N* = 56 137 |  | *N* = 44 637 |  |
|  | count | (%) | count | (%) |
| **Age, years** |  |  |  |  |
| 0–14 | 98 | (0.2%) | .. | .. |
| 15–29 | 3 121 | (5.6%) | 374 | (0.8%) |
| 30–39 | 13 963 | (24.9%) | 6 681 | (15.0%) |
| 40–49 | 17 816 | (31.7%) | 15 963 | (35.8%) |
| 50–59 | 13 941 | (24.8%) | 8 028 | (18.0%) |
| 60+ | 7 198 | (12.8%) | 13 422 | (30.1%) |
| Unknown | .. | .. | 169 | .. |
| **Gender** |  |  |  |  |
| Female | .. | .. | 13 084 | (29.4%) |
| Male | .. | .. | 31 427 | (70.6%) |
| Transgender | .. | .. | 10 | (<0.1%) |
| Unknown | .. | .. | 116 | .. |
| **Ethnicity** |  |  |  |  |
| Asian | .. | .. | 3 400 | (8.4%) |
| Black | .. | .. | 1 468 | (3.6%) |
| White | .. | .. | 33 721 | (83.5%) |
| Mixed | .. | .. | 445 | (1.1%) |
| Other | .. | .. | 1 434 | (3.6%) |
| Unknown | .. | .. | 4 169 | .. |
| **Drug Use** |  |  |  |  |
| Current | .. | .. | 11 742 | (33.8%) |
| Past | .. | .. | 13 869 | (39.9%) |
| Never | .. | .. | 9 083 | (26.1%) |
| Unknown | .. | .. | 9 943 | .. |
| **Disease Stage** |  |  |  |  |
| Decompensated cirrhosis | 1 608 | (3.4%) | 1 010 | (2.8%) |
| Compensated cirrhosis | 9 214 | (19.5%) | 7 554 | (20.7%) |
| Unspecified cirrhosis | 171 | (0.4%) | .. | .. |
| No cirrhosis | 44 900 | (80.3%) | 35 485 | (76.4%) |
| Unknown | 244 | .. | 588 | .. |
| **Treatment History** |  |  |  |  |
| Relapsed | 26 | (<0.1%) | .. | .. |
| Treatment Experienced | 6 807 | (13.5%) | .. | .. |
| Treatment Naïve | 43 717 | (86.5%) | .. | .. |
| Unknown | 5 587 | .. | .. | .. |

The time-series of HCV cases identified, SVRs and drug service referrals for each ODN are shown in Figures 1(a)-(c). Outcomes varied widely across ODNs. 4,852 HCV-infected individuals were identified in ‘Birmingham’, the largest ODN; while 1,075 HCV-infected were identified in ‘Surrey’, the smallest ODN. In ‘Birmingham’, 2,494 SVRs, and 1,092 drug service referrals were observed. The ODN ‘South Thames’ saw 2,756 SVRs during the study period, more than ‘Birmingham’, owing to a higher treatment success rate. In ‘Surrey’, only 673 SVRs, and 126 drug service referrals were observed. During the first English COVID-19 lockdown (March - May 2020) all the outcomes fell sharply across all ODNs.

**Figure 1:**
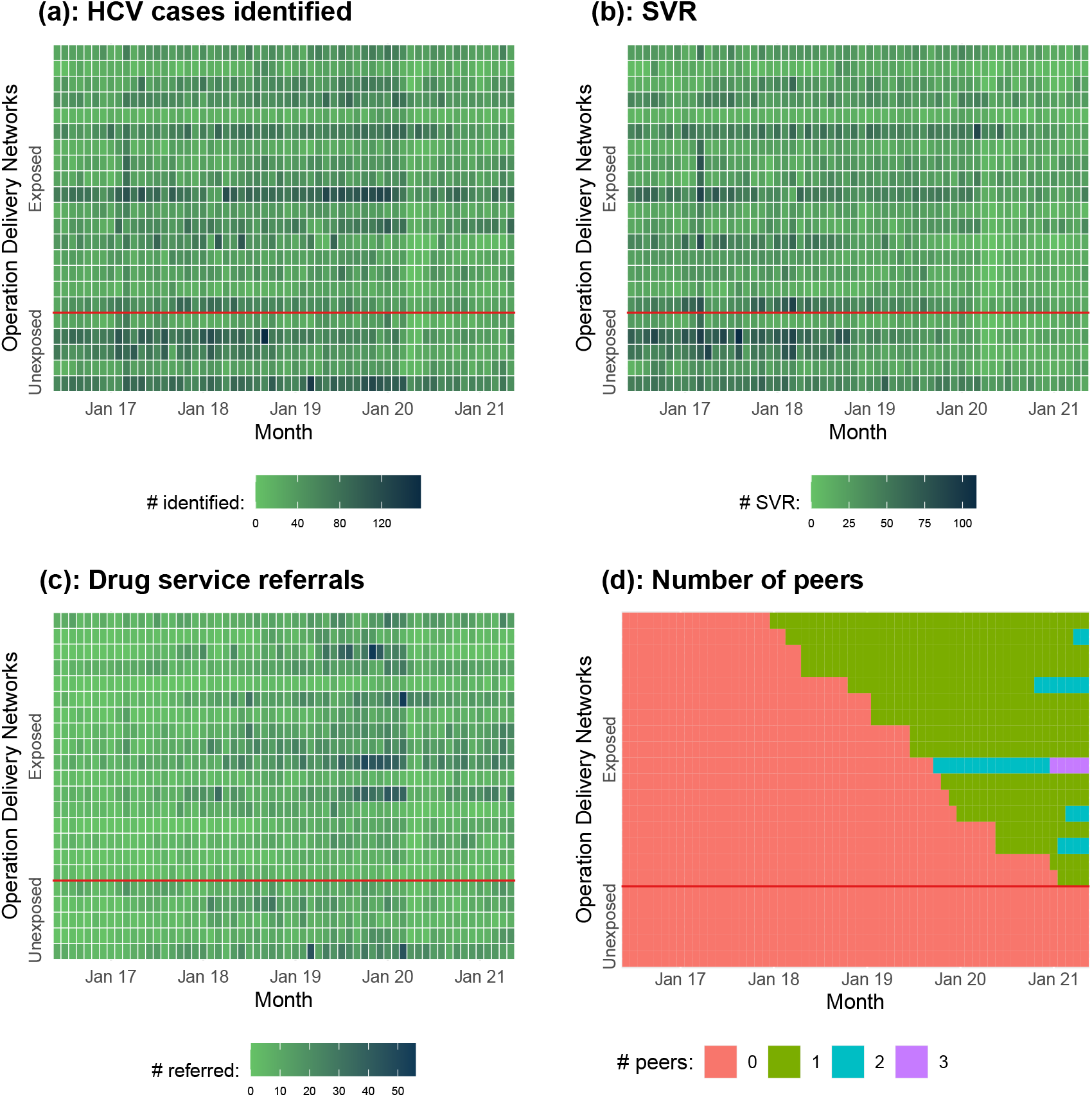
Graphical summaries of outcomes and exposure. Data are shown for each Operation Delivery Network and each month between June 2016 and May 2021: (a) number of Hepatitis C virus (HCV) infected individuals identified (Blueteq dataset); (b) number of individuals completing DDA treatment and achieving confirmed sustained viral response (Registry); (c) the number of individuals initiating DDA treatment referred from drug services (Registry); (d) number of peers (HCT data). Abbreviations: Jan: January; #: Number.

Figure 1(d) displays the number of peers in each ODN between June 2016 and May 2021. The first peer began working in the ODN ‘Nottingham’ in January 2018. By the end of the study period, five ODNs remained without a peer, five ODNs employed more than one peer, and there were 23 peers operating nationally.

Table 2 summarises characteristics of individuals supported by peers, including year of first contact, their gender, setting of first engagement, injecting and homelessness status, and whether they were clients of drug services. The data included 18,999 individuals. A large share of individuals recorded in the HCT data base belonged to groups traditionally underserved by HCV health services, including those currently (1,620 [34·6% of non-missing cases]) or previously (2,748 [58·6% of non-missing cases]) injecting drugs, and those experiencing homelessness (1,582 [17·6% of non-missing cases]).

**Table 2:**
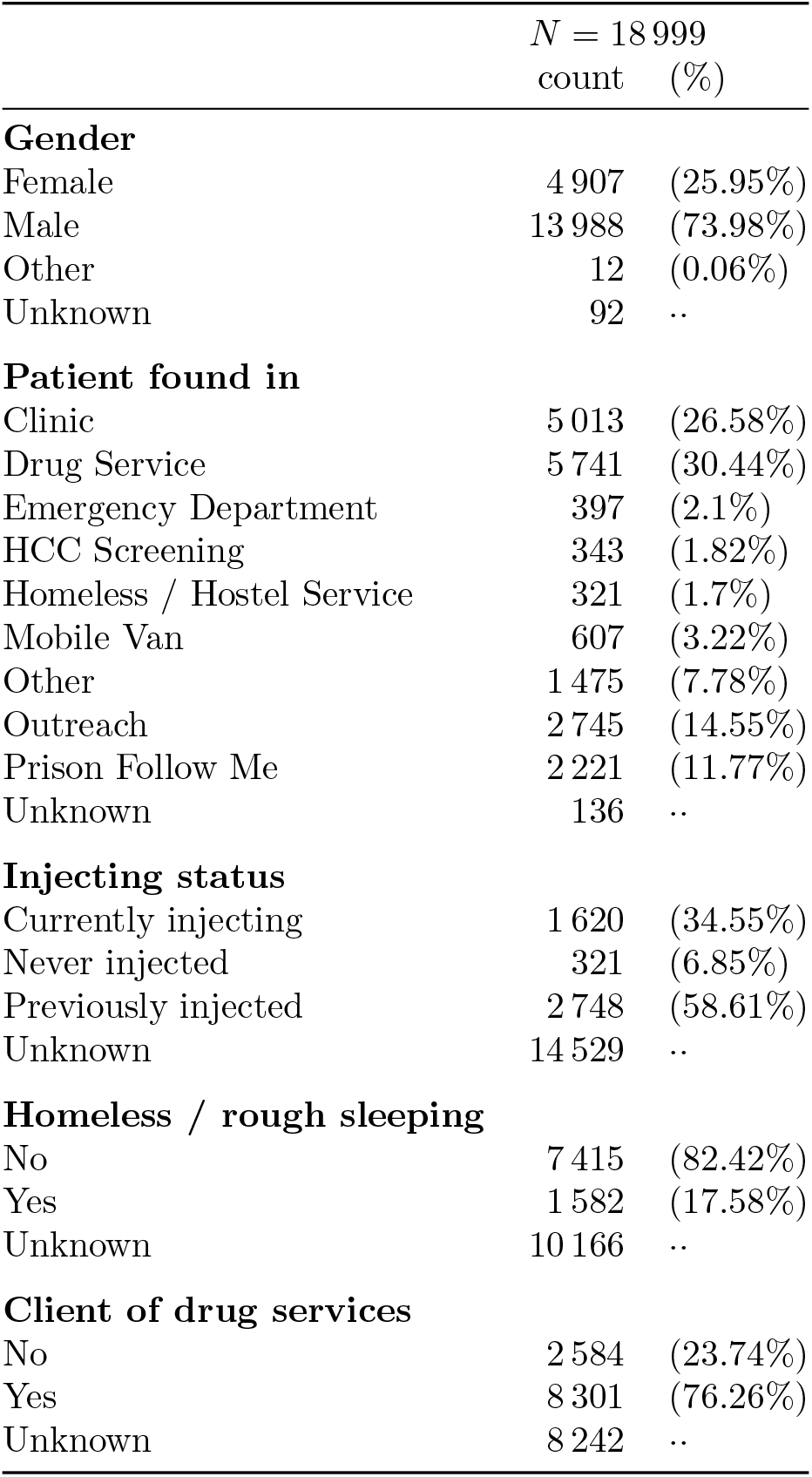
Characteristics of individuals supported by peers. Results are shown as count (%). Characteristics are for individuals recorded the HCT database who started treatment between January 2017 and December 2024. Table 6 in Appendix C shows the number of individuals supported by peers each year. Percentages were calculated using non-missing observations.

| | $N = 18\,999$ | |
| --- | --- | --- |
|  | count | (%) |
| **Gender** |  |  |
| Female | 4 907 | (25.95%) |
| Male | 13 988 | (73.98%) |
| Other | 12 | (0.06%) |
| Unknown | 92 | .. |
| **Patient found in** |  |  |
| Clinic | 5 013 | (26.58%) |
| Drug Service | 5 741 | (30.44%) |
| Emergency Department | 397 | (2.1%) |
| HCC Screening | 343 | (1.82%) |
| Homeless / Hostel Service | 321 | (1.7%) |
| Mobile Van | 607 | (3.22%) |
| Other | 1 475 | (7.78%) |
| Outreach | 2 745 | (14.55%) |
| Prison Follow Me | 2 221 | (11.77%) |
| Unknown | 136 | .. |
| **Injecting status** |  |  |
| Currently injecting | 1 620 | (34.55%) |
| Never injected | 321 | (6.85%) |
| Previously injected | 2 748 | (58.61%) |
| Unknown | 14 529 | .. |
| **Homeless / rough sleeping** |  |  |
| No | 7 415 | (82.42%) |
| Yes | 1 582 | (17.58%) |
| Unknown | 10 166 | .. |
| **Client of drug services** |  |  |
| No | 2 584 | (23.74%) |
| Yes | 8 301 | (76.26%) |
| Unknown | 8 242 | .. |

### Statistical analysis

Employment of peers increased the number of HCV cases identified by 13·9% (95% CrI [5·3, 21·7]; PPos=99·8%) across all post-intervention periods compared to the counterfactual of no peer intervention (Figure 2). This corresponded to an overall increase in case-finding of 2166 individuals (95% CrI [828, 3387]; Figure 6 in Appendix C). There was some evidence that the peers intervention increased overall SVRs (PPos=91·1%) and drug services referrals (PPos=84·2%; Figure 2). The positive effects of the peers intervention were more pronounced during the first national COVID-19 lockdown. We estimated that HCV-cases indentified increased by 34·8% (95% CrI [19.4, 48·1], PPos*>*99·9%), SVRs increased by 28·2% (95% CrI [4·0, 46·3], PPos=98·7%), and drug service referrals increased by 31·1% (95% CrI [6·4, 50·8], PPos=99·3%) due to the work of peers (Figure 2).

**Figure 2:**
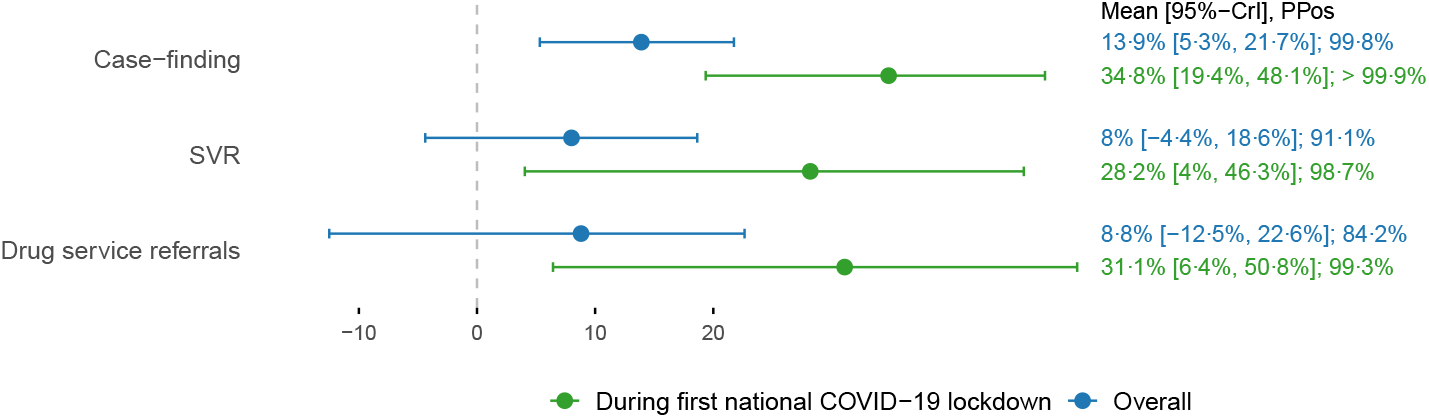
Mean and 95% credible intervals of the percentage change in each outcome due to the peers intervention. Outcomes considered were: the number of HCV cases identified, the number of sustained viral responses, and the number of drug service referrals. The percentage change in each outcome was calculated across all post-intervention time periods (blue) and post-intervention time periods during with the first COVID-19 lockdown was in place (green). For a detailed definition of outcomes and the model see Appendices B and D. HCV = Hepatitis C virus; SVR = sustained viral response.

The RRs for the number of HCV cases identified exhibited a non-linear relationship with cumulative peer-months (Figure 3 [a]). At low exposure intensities (*<*eight peer-months), there was no evidence of an intervention effect, though the RR rose slowly with increasing intensity. At moderate intensities (8-18 peer-months), RRs rose sharply with exposure intensity and almost certainly exceed 1·1. Finally, at high intensities (*>*18 peer-months), the RRs slightly decreased, but remained larger than one with very high probability. Similar patterns were observed for RR for SVRs and drug services referrals (Figures 3 (b) and (c)) although for high intensities the 95% CrIs included zero.

**Figure 3:**
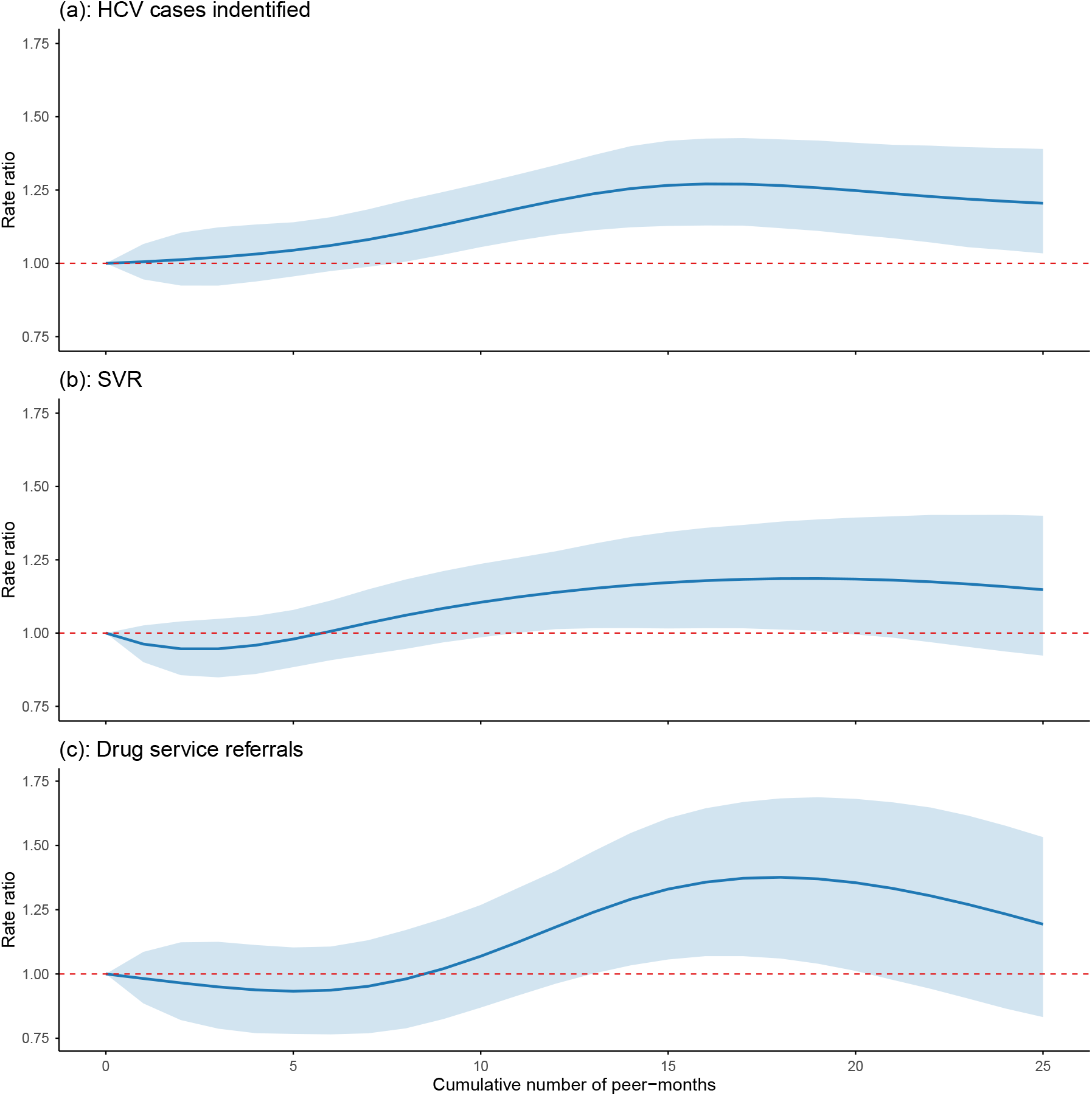
Means and 95% credible intervals of rate ratios by exposure intensity. Outcome considered were: the number of HCV cases identified; the number of sustained viral responses; and the number of drug service referrals when the cumulative number of peer-months served as a measure of exposure intensity. HCV = Hepatitis C virus; SVR = sustained viral response.

## 4 Discussion

In this work we set out to improve and extend the evidence on the effect of peers on HCV-treatment related outcomes in England between January 2016 and May 2021. We found strong evidence that peer support increased the number of HCV-infected individuals identified estimating a 14% increase in case-finding (≈ 2200 HCV-infected individuals, PPos=99·8%). Our analyses further suggested that the peer intervention may have had a positive impact on SVRs (8.0% increase, PPos 91·1%) and drug service referrals (8·8% increase, PPos 84·42%).

Our findings on case-funding are particularly important as they indicate that peers are able to reach out to HCV-infected individuals that would not have been identified by other NHS services. As shown by the HCT patient database (Table 2), many of the individuals supported by peers belonged to marginalised groups with limited access to healthcare.

We further found that the positive effects of the peer intervention were particularly pronounced during the first English COVID-19 lockdown. This might reflect that peer support was one of the few available routes to HCV care after staff redeployment and distancing measures limited capacity of clinical services. Peers were also actively involved in an initiative providing HCV testing and treatment to homeless individuals who had been given temporary accommodation throughout the pandemic and were thus easier to engage during this period. ^29,30^

The effect of the peers intervention varied with exposure intensity, quantified by the number of peer-months of exposure. During the first few (roughly eight) peer-months, there was no significant evidence of an effect. This likely reflects the time required for peers to train and establish networks within their ODNs. The effect strengthened and increased with exposure intensity between roughly eight and 18 peer-months and plateaued at high intensity (*>*20 peer-months). A possible explanation is that, as individuals who are more open to treatment are recruited, the remaining population becomes increasingly hard to identify and engage.

Our study has several strengths. First, it was the first to look at the effect of peer support workers on the number of HCV cases identified. Second, we exploited the staggered adoption of the peers intervention as a natural quasi-experiment using a state-of-the-art Bayesian causal factor analysis approach. This allowed us to draw inferences regarding the effectiveness of peers without the need for a randomised trial. Third, to the best of our knowledge, our study was the first to model heterogeneity in the effects of peer support and to identify some of its drivers, including exposure intensity and the presence of lockdowns. Fourth, by analysing data over a time period up to 18 months after the first peer started working, we were able to assess the long-term effects of peer support. This is important because, as shown in our analysis, some time is required before the effects of the intervention become evident. Fifth, using HCT data, we demonstrated that many of the individuals supported by peers belonged to marginalised population groups.

We acknowledge several limitations. First, we did not assess the effect of the peers intervention beyond May 2021. This would be challenging because there were very few unexposed ODNs left for comparison. Second, we did not examine some other outcomes of interest, including deaths due to HCV, number of reinfections, and sustained viral clearances at six months. Estimating the effect of the intervention on these outcomes would be hard, as they involve large amounts of missing data and low counts, which limit statistical power. A potential solution could be to model these outcomes jointly with existing outcomes to improve efficiency. Third, the HCT data had a high proportion of missing data in some variables.

Our study has important policy implications. We provided evidence that peer support improved multiple epidemiological outcomes, highlighting the need to sustain this intervention to achieve the WHO HCV elimination targets. Our analyses suggested that the effect of peers was more pronounced during the COVID-19 lockdown. Thus, recruitment of peer supporters may be an effective strategy for maintaining patient engagement during future periods when health services cannot operate at full capacity and/or barriers to health care access are particularly high. We also showed that many of the individuals supported by peers are from marginalised populations that tend to be excluded from traditional health services. Finally, since the effect of peer support appeared to have a complex, non-linear relationship with intervention intensity, an economic evaluation to identify optimal rollout strategies in other countries and/or disease areas is warranted.

## Patient and Public Involvement statement

As this study represents a retrospective evaluation, it was not possible to involve patients in the design of the intervention. Research questions were developed with in correspondence HCT staff, who have relevant lived experience.

## Ethics statement

The analyses are based on a retrospective review of fully anonymised data collected as part of routine clinical care and disease surveillance on an opt-out basis. No identifiable patient data were used.

In accordance with guidance from the Health Research Authority (https://www.hra.nhs.uk/planning-and-improving-research/policies-standards-legislation/data-protection-and-information-governance/gdpr-guidance/), this work does not constitute research requiring review by an NHS Research Ethics Committee and therefore did not require formal ethics approval. Appropriate governance permissions for use of the registry data were in place.

## Acknowledgments

ChatGPT (model GPT-5.1) operated by OpenAI was used as a thessaurus. The prompt was ‘What is a synonym for …’. ChatGPT was also used to improve the quality of already existing graphs. One would input the code for an existing graph (never the data) and prompt a specific request like: ‘Increase the font size on the x-axis’.

## Data availability statement

Data can be obtained from the authors upon request for research purposes. Code will be made available in GitHub.

## A Search strategy

To build an understanding of the available evidence on the effect of peers support workers on HCV related treatment outcomes, we updated a recent review paper by Li and colleagues. ^31^ The original review included papers between 1 January 2010 and 12 March 2025. We included papers published up to 30 November 2025. We considered publications listed on PubMed using the following search terms:

((hepatitis[Title/Abstract] OR hepadnaviridae[Title/Abstract] OR hep-b[Title/Abstract] OR hep-c[Title/Abstract] OR hep-d[Title/Abstract] OR hepatitides[Title/Abstract] OR HBV[Title/Abstract] OR HCV[Title/Abstract] OR HDV[Title/Abstract]) AND (community-based[Title/Abstract] OR community[Title/Abstract] OR communities[Title/Abstract] OR communal[Title/Abstract] OR communalism[Title/Abstract] OR communally[Title/Abstract] OR commune[Title/Abstract] OR local[Title/Abstract] OR localisation[Title/Abstract] OR localise[Title/Abstract] OR localised[Title/Abstract] OR localises[Title/Abstract] OR localising[Title/Abstract] OR localization[Title/Abstract] OR localize[Title/Abstract] OR localized[Title/Abstract] OR localizes[Title/Abstract] OR localizing[Title/Abstract] OR locally[Title/Abstract] OR town[Title/Abstract] OR grassroots[Title/Abstract] OR grassroot[Title/Abstract] OR “grass root”[Title/Abstract] OR “grass roots”[Title/Abstract] OR “around town”[Title/Abstract] OR people-centered[Title/Abstract] OR people-centred[Title/Abstract] OR patient-centered[Title/Abstract] OR patient-centred[Title/Abstract] OR person-centered[Title/Abstract] OR person-centred[Title/Abstract] OR lay[Title/Abstract] OR public[Title/Abstract] OR public’s[Title/Abstract] OR publically[Title/Abstract] OR publics[Title/Abstract] OR peer[Title/Abstract] OR personally-tailored[Title/Abstract] OR individually-tailored[Title/Abstract] OR individual[Title/Abstract] OR individual’s[Title/Abstract] OR individualisation[Title/Abstract] OR individualise[Title/Abstract] OR individualised[Title/Abstract] OR individualising[Title/Abstract] OR individualities[Title/Abstract] OR individuality[Title/Abstract] OR individualization[Title/Abstract] OR individualize[Title/Abstract] OR individualized[Title/Abstract] OR individualizes[Title/Abstract] OR individualizing[Title/Abstract] OR individually[Title/Abstract] OR individuals[Title/Abstract] OR individuate[Title/Abstract] OR individuated[Title/Abstract] OR individuates[Title/Abstract] OR individuating[Title/Abstract] OR individuation[Title/Abstract] OR persons[Title/Abstract] OR “shared decision making”[Title/Abstract] OR stakeholder[Title/Abstract] OR stakeholder’s[Title/Abstract] OR stakeholders[Title/Abstract] OR consumers[Title/Abstract] OR consume[Title/Abstract] OR consumed[Title/Abstract] OR consumer[Title/Abstract] OR consumer’s[Title/Abstract] OR consumes[Title/Abstract] OR consuming[Title/Abstract] OR co-design[Title/Abstract] OR advocacy[Title/Abstract] OR advocacies[Title/Abstract] OR advocacy’s[Title/Abstract] OR advocate[Title/Abstract] OR advocating[Title/Abstract] OR advocate’s[Title/Abstract] OR advocated[Title/Abstract] OR advocates[Title/Abstract] OR advocation[Title/Abstract] OR communication[Title/Abstract] OR communicate[Title/Abstract] OR communicated[Title/Abstract] OR communicates[Title/Abstract] OR communicating[Title/Abstract] OR communicational[Title/Abstract] OR communicatively[Title/Abstract] OR communicativeness[Title/Abstract] OR communicator[Title/Abstract] OR communicator’s[Title/Abstract] OR communicators[Title/Abstract] OR engage[Title/Abstract] OR engaged[Title/Abstract] OR engagement’s[Title/Abstract] OR engagements[Title/Abstract] OR engages[Title/Abstract] OR engaging[Title/Abstract] OR “social participation”[ Title/Abstract] OR engagement[Title/Abstract] OR participation[Title/Abstract] OR participant[Title/Abstract] OR participant’s[Title/Abstract] OR participants[Title/Abstract] OR participate[Title/Abstract] OR participated[Title/Abstract] OR participates[Title/Abstract] OR participating[Title/Abstract] OR participations[Title/Abstract] OR participative[Title/Abstract] OR participator[Title/Abstract] OR participators[Title/Abstract] OR involvement[Title/Abstract] OR involve[Title/Abstract] OR involved[Title/Abstract] OR involvements[Title/Abstract] OR involves[Title/Abstract] OR involving[Title/Abstract] OR messaging[Title/Abstract] OR message[Title/Abstract] OR message’s[Title/Abstract] OR messaged[Title/Abstract] OR messages[Title/Abstract] OR storytelling[Title/Abstract] OR storyteller[Title/Abstract] OR storyteller’s[Title/Abstract] OR storytellers[Title/Abstract] OR story[Title/Abstract] OR stories[Title/Abstract] OR empowerment[Title/Abstract] OR navigation[Title/Abstract] OR navigate[Title/Abstract] OR navigated[Title/Abstract] OR navigates[Title/Abstract] OR navigating[Title/Abstract] OR navigations[Title/Abstract] OR navigator[Title/Abstract] OR navigator’s[Title/Abstract] OR navigators[Title/Abstract] OR contribution[Title/Abstract] OR con-tribute[Title/Abstract] OR contributed[Title/Abstract] OR contributes[Title/Abstract] OR contributing[Title/Abstract] OR preference[Title/Abstract] OR preferences[Title/Abstract])) AND (intervention[Title/Abstract] OR experiment[Title/Abstract] OR trial[Title/Abstract] OR campaign[Title/Abstract] OR implementation[Title/Abstract] OR method[Title/Abstract]) NOT (acetylcysteine[Title] OR adefovir[Title] OR lamivudine[Title] OR ribavirin[Title] OR interferon[Title] OR peginterferon[Title] OR “ursodeoxycholic acid”[Title] OR baraclude[Title] OR telbivudine[Title] OR entecavir[Title] OR tenofovir[Title] OR elbasvir[Title] OR grazoprevir[Title] OR glecaprevir[Title] OR pentoxifylline[Title] OR pibrentasvir[Title] OR pradefovir[Title] OR ravidasvir[Title] OR sofosbuvir[Title] OR tacrolimus[Title] OR telaprevir[Title] OR velpatasvir[Title] OR rituximab[Title] OR Durvalumab[Title] OR placebo[Title] OR “systematic review”[Title] OR “phase 1”[Title] OR “phase I”[Title] OR “phase 2”[Title] OR “phase II”[Title] OR “phase 3”[Title] OR “phase III”[Title] OR “phase 4”[Title] OR “phase IV”[Title] OR “case report”[Title] OR “in vivo”[Title] OR “in vitro”[Title] OR immunogenicity[Title] OR “efficacy and safety”[Title] OR “safety, efficacy”[Title] OR “efficacy, safety”[Title])

PubMed filters applied: text availability: Free Full Text, Clinical Study, Clinical Trial, Controlled Clinical Trial, Observational Study, Randomized Controlled Trial, Humans, English.

We then did a Google Scholar forward search for each of the papers identified. We used the following inclusion and exclusion criteria throughout:

**Table 3:** Inclusion and exclusion criteria of systematic review.

|  |  |
| --- | --- |
| Inclusion criteria | 1. Report on at least one outcome in the hepatitis C care continuum (testing, diagnosis, treatment, or cure)<br>2. Studies that have a comparator group (including randomized controlled trials and observational studies)<br>3. Chronic viral HCV<br>4. English language<br>5. Sufficient description of community engagement activities that enable us to assess and define the level of engagement<br>6. Primary outcome data available<br>7. Compare standard of care plus peer support (intervention) versus standard of care (control) |
| Exclusion criteria | 1. Study subjects were fewer than five<br>2. Overlap of study population or results with another included publication<br>3. Interventions that did not have a comparison or control arm<br>4. No or insufficient information about community engagement to determine the level of engagement, or the intervention was non-participatory and/or manipulative (e.g., viral hepatitis treatment and therapy)<br>5. Focused on other types of hepatitis diseases (e.g., hepatitis A and E or alcoholic hepatitis) |

Inclusion criteria 1. and 3. were tightened compared to Li et al’s review, which considered viral hepatitis (hepatitis B, C, or D). Inclusion criteria 7. was added for the purpose of this study and was not part of the inclusion criteria in Li et al’s review. Studies with multiple intervention conditions one of which was standard of care plus peer support were included. Studies that do not consider peers, include peers as part of a larger intervention (and thus the effect of peers alone cannot be isolated), or employ peers in all intervention arms were not included.

## B Data cleaning details and detailed description of outcomes

**Figure 4:**
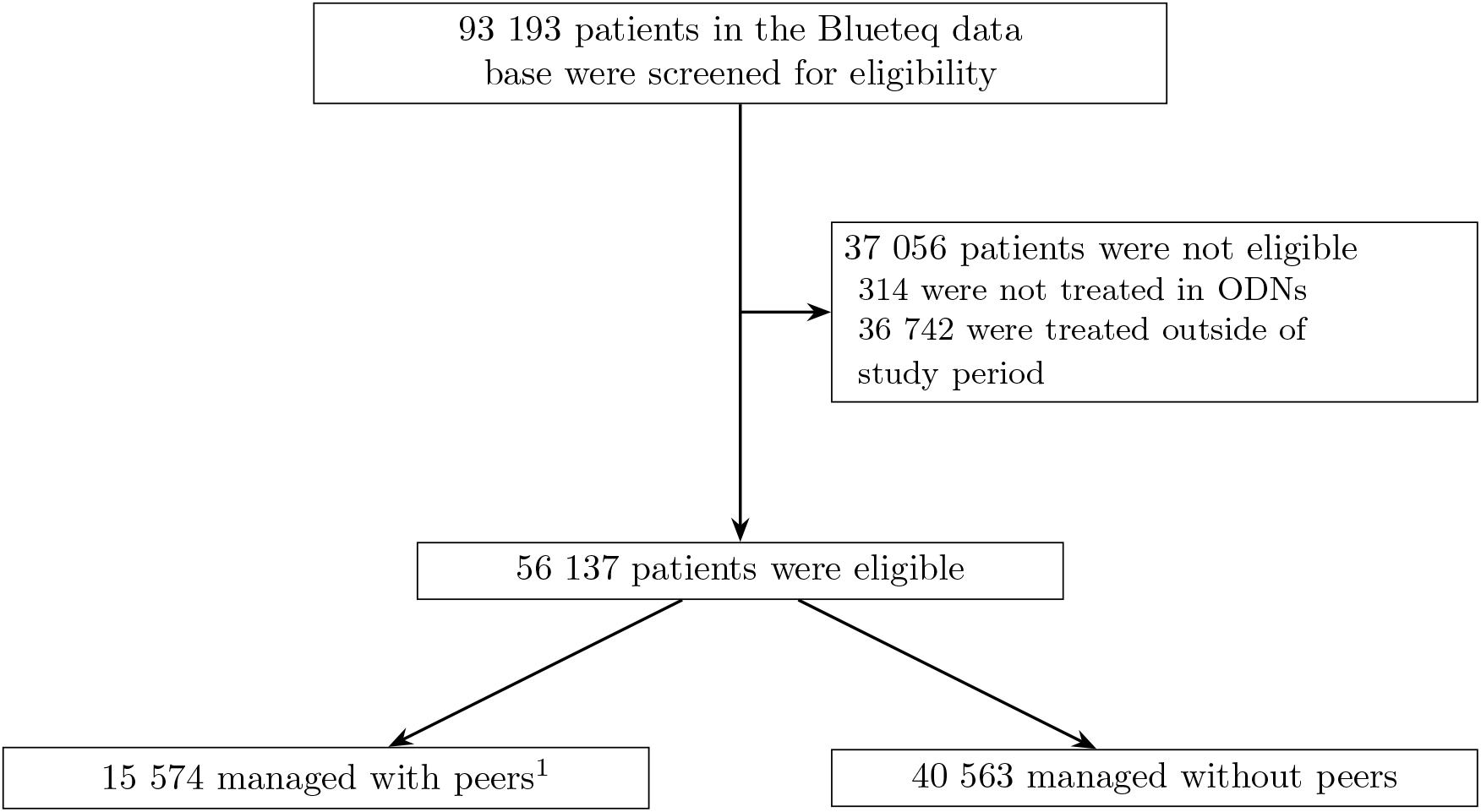
Profile of patients in Blueteq data base. ODN: Operation delivery network. ^1^Patients were considered ‘managed with peers’ if peers were active in the ODN while the patient was treated.

**Table 4:** Definition of primary and secondary outcomes. DAA = direct-acting antivirals; HCV = Hepatitis C virus; SVR = sustained viral response.

| Outcome | Explanation |
| --- | --- |
| HCV cases identified | The number of individual treatment funding requests, extracted from Blueteq. Since all HCV infected individuals are DAA treatment eligible, once a patient’s HCV infection has been confirmed, an individual treatment funding request is put into Blueteq, so DDA treatments can be ordered. Without approved funding request, providers are not reimbursed for treatment. Thus, Blueteq is a complete data base of all HCV-infected individuals identified by NHSE. We use the date the individual treatment funding request was put into Blueteq as the date the individual was confirmed as HCV-infected. |
| SVR | The number of patients recorded as having completed the full course of DAA medication and having achieved a confirmed sustained viral response (SVR, clearance of HCV from their body), extracted from the Registry. HCV clearance was confirmed at a follow-up meeting 12 weeks after treatment completion. The date of the first appointment with HCV services was used as the relevant date when a patient initiated successful treatment. |
| Drug service referrals | The number of patients recorded as initiating HCV treatment who were referred by drug services, extracted from the Registry. A patient was considered to have initiated treatment once they were assigned a date for their first appointment with HCV treatment services. The date of the first appointment was used as the treatment initiation date. |

**Figure 5:**
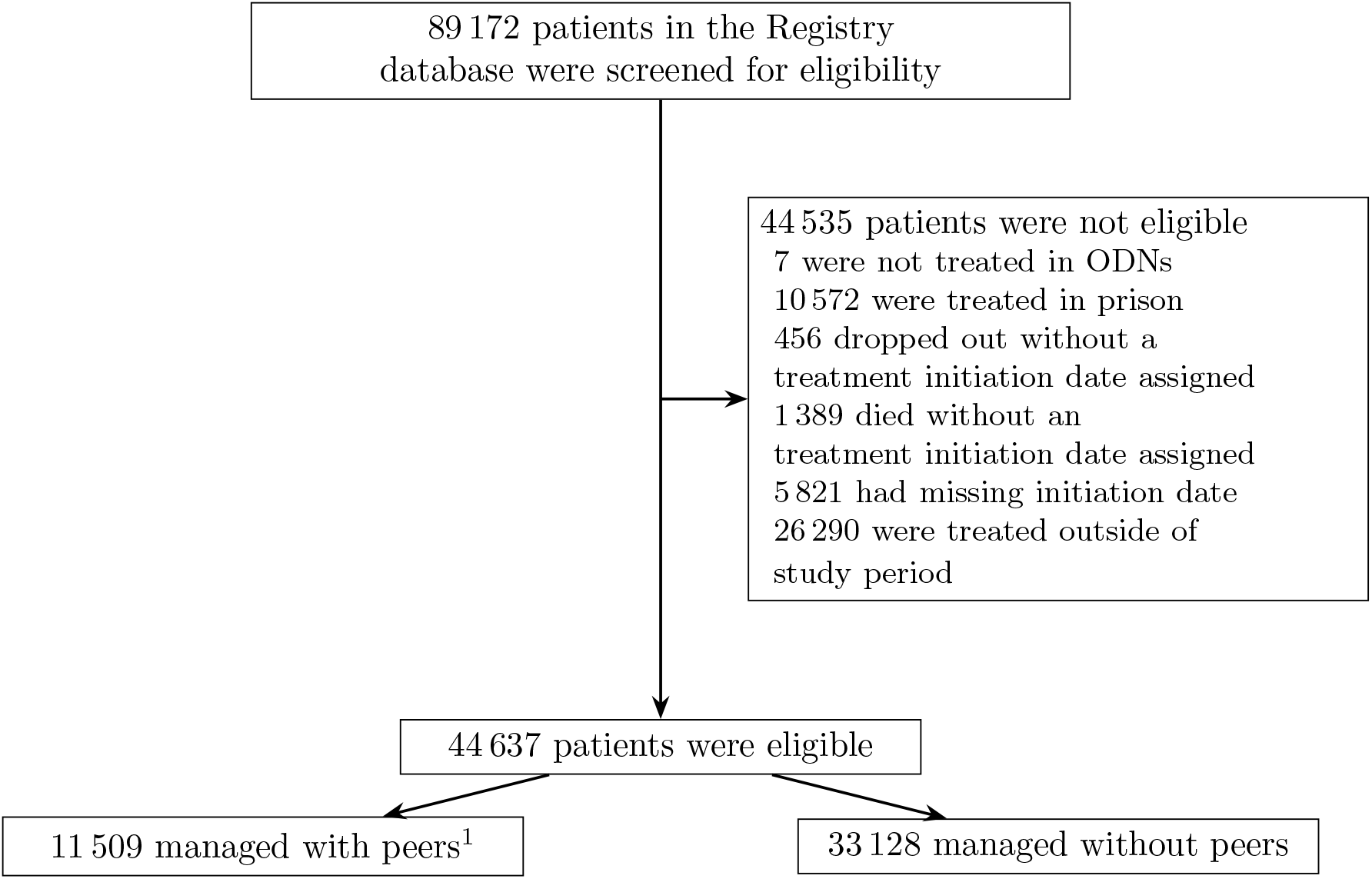
Profile of patients in the English national hepatitis C treatment registry. ODN: Operational delivery network. ^1^Patients were considered ‘managed with peers’ if peers were active in the ODN while the patient was treated.

## C Additional results

**Table 5:** Additional characteristics of study sample. Results shown are count (%). Percentages are calculated using non-missing observations. Characteristics and treatment outcomes are for HCV-infected individuals recorded in the English national Hepatitis C treatment registry between June 2016 and May 2021. HCV = Hepatitis C virus, GP = general practitioner. ^1^ Non-response: Remained HCV RNA positive and did not clear the virus throughout treatment or post-treatment; Relapse: cleared during treatment but became positive post-treatment; Breakthrough: cleared during treatment but became positive again during treatment. ^2^ HCV RNA negative 12 weeks after completion of treatment.

|  | Count (%) |
| --- | --- |
| **Outcome** |  |
| Death before completion | 792 (1.8%) |
| Lost to follow-up | 5,295 (12.3%) |
| Non-response/Relapse/Breakthrough<sup>1</sup> | 1,787 (4.1%) |
| Other | 465 (1.1%) |
| Sustained viral response<sup>2</sup> | 34,813 (80.7%) |
| Unknown | 1,485 (·) |
| **Likely Route of Transmission** |  |
| Healthcare (e.g., organs, blood) | 2,884 (9.3%) |
| Mother to child | 234 (0.8%) |
| Occupational contact | 149 (0.5%) |
| Other blood exposure (e.g., tattoo) | 1,490 (4.8%) |
| PWID | 24,673 (79.4%) |
| Sexual | 1,656 (5.3%) |
| Unknown | 13,551 (·) |
| **HCV Genotype** |  |
| Genotype 1 | 21,183 (47.6%) |
| Genotype 2 | 2,198 (4.9%) |
| Genotype 3 | 17,108 (38.4%) |
| Other | 4,057 (9.1%) |
| Unknown | 91 (·) |
| **Source of Referral** |  |
| Antenatal | 156 (0.4%) |
| Drug services | 11,513 (26.0%) |
| GP or Pharmacy | 16,482 (37.2%) |
| Genitourinary Medicine | 1,707 (3.9%) |
| Health Outreach | 764 (1.7%) |
| Hospital | 10,471 (23.7%) |
| Psychiatry | 250 (0.6%) |
| Self-referral | 187 (0.4%) |
| Other | 2,738 (6.2%) |
| Unknown | 369 (·) |

**Table 6:**
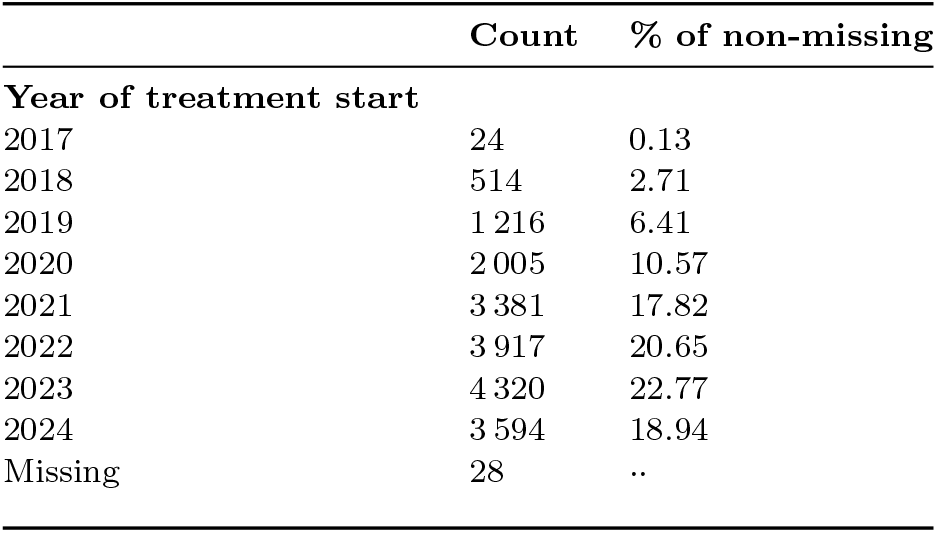
Number of HCV-infected individuals supported by peers by year. Peer support may start any time while an individual is in Hepatitis C virus treatment. This explains why some individuals who started treatment before 2018 received peer support. HCV = Hepatitis C virus.

**Figure 6:**
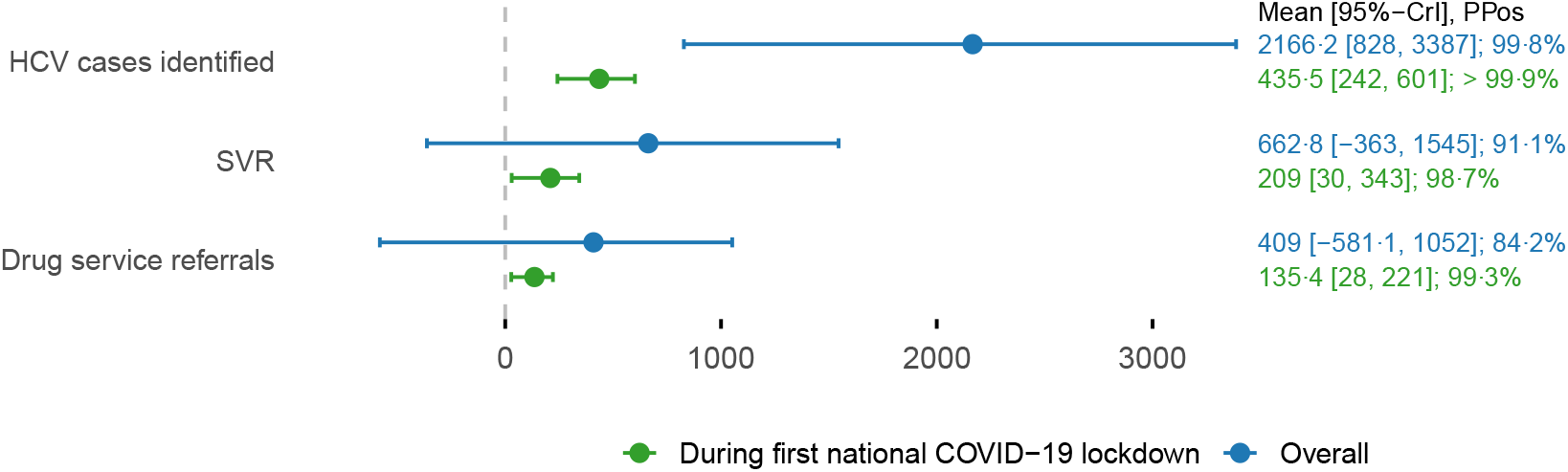
Mean and 95% credible intervals of the change due to the peers intervention. Outcomes considered were: number of HCV cases identified; number of sustained viral responses; and number of drug service referrals. The change in each outcome was calculated across all post-intervention time periods (blue) and post-intervention time periods during with the first COVID-19 lockdown was in place (green). HCV = Hepatitis C virus; SVR = sustained viral response.

**Figure 7:**
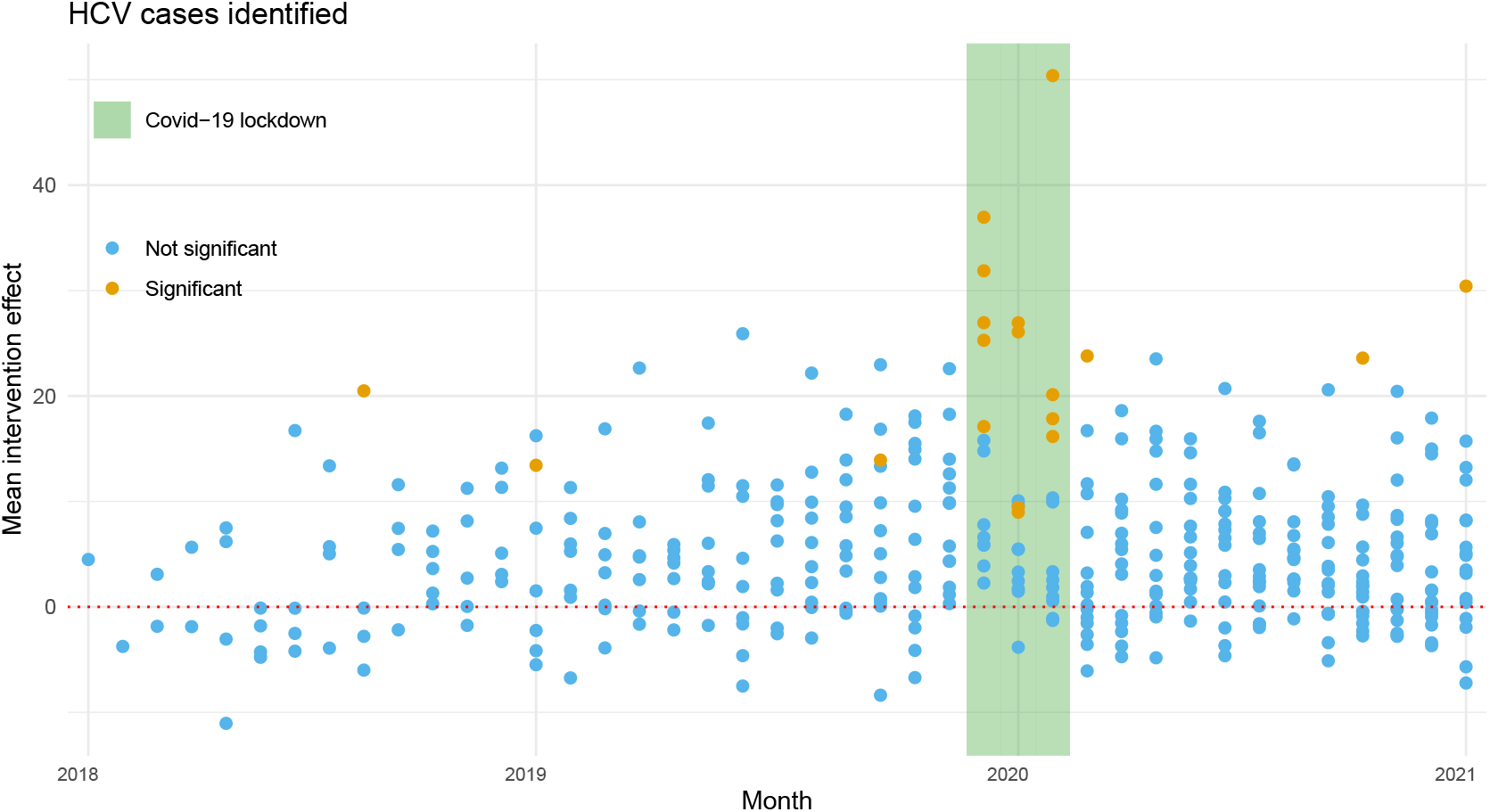
Individual intervention effects for HCV cases identified. Individual intervention effects were defined as the difference between the observed outcome and the intervention-free counterfactual. The graph shows posterior means of individual intervention effects for each ODN during each post-intervention month based on the Bayesian causal factor model. HCV = Hepatitis C virus.

**Figure 8:**
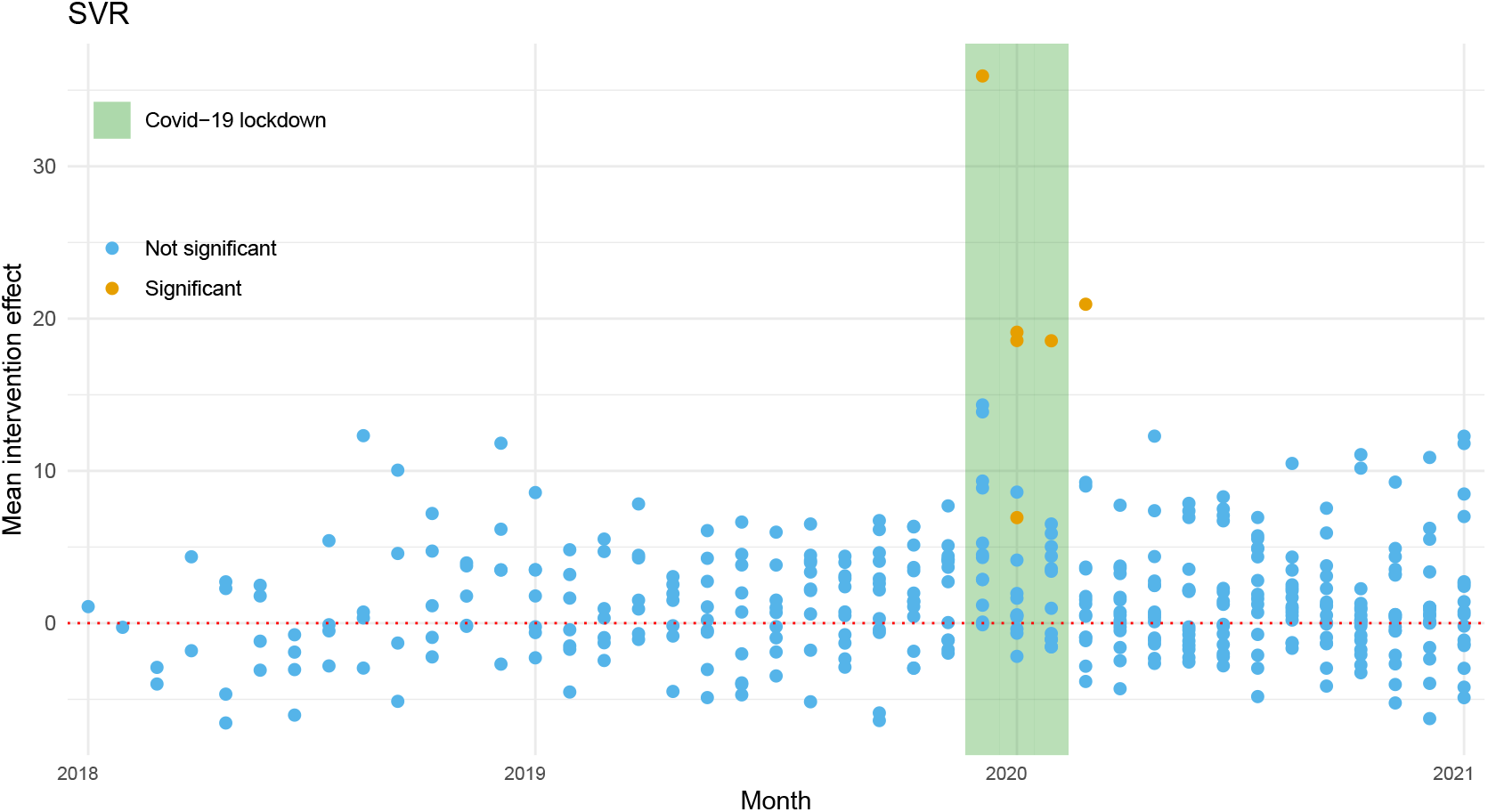
Individual intervention effects for SVR. Individual intervention effects were defined as the difference between the observed outcome and the intervention-free counterfactual. The graph shows posterior means of individual intervention effects for each ODN during each post-intervention month based on the Bayesian causal factor model. SVR = sustained viral response.

**Figure 9:**
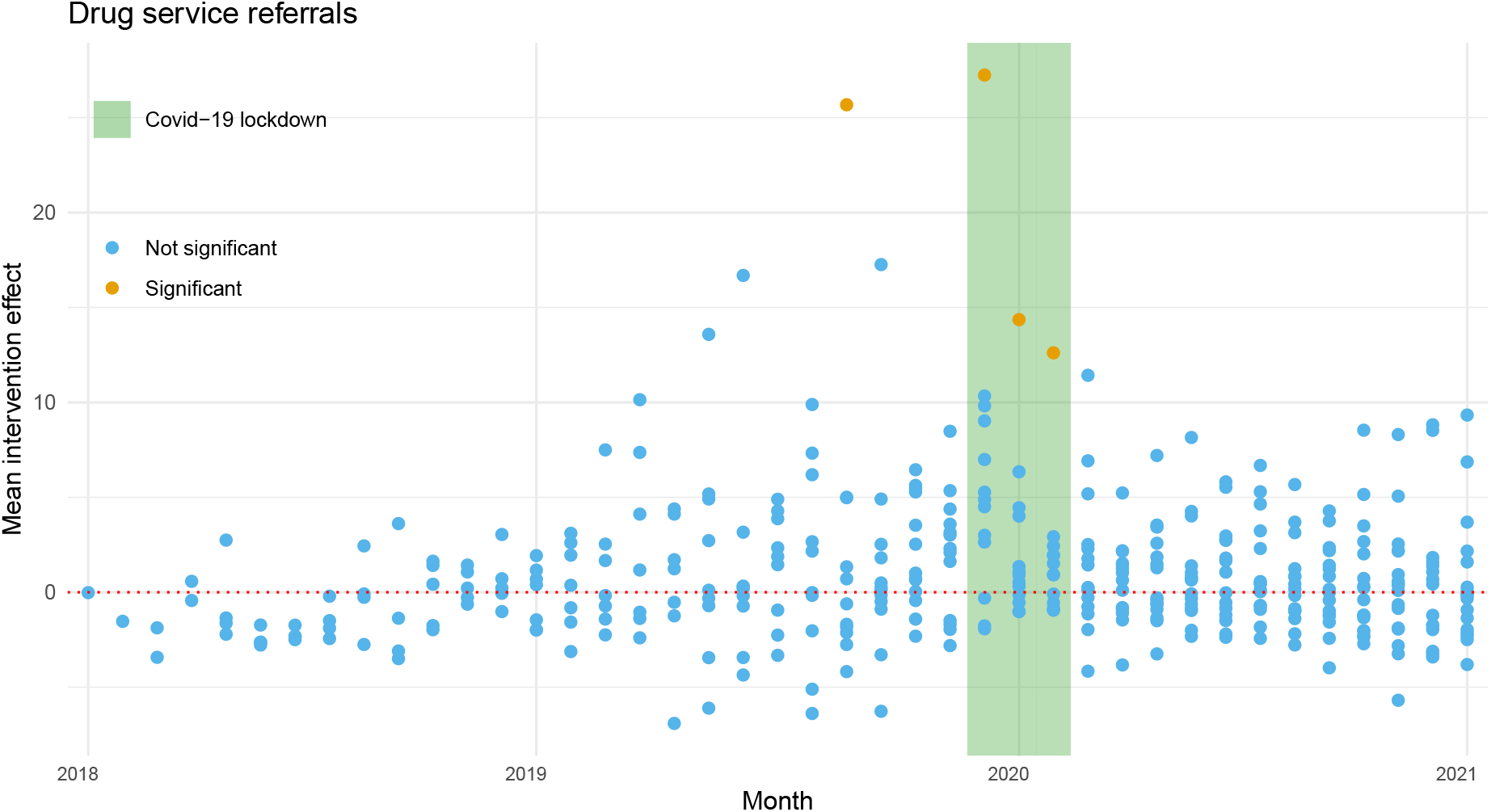
Individual intervention effects for drug service referrals. Individual intervention effects were defined as the difference between the observed outcome and the intervention-free counterfactual. The graph shows posterior means of individual intervention effects for each ODN during each post-intervention month based on the Bayesian causal factor model.

## D Details of statistical modelling

In this section, we outline the statistical methodology that we used to evaluate the peers intervention. For full details, see Schmidt and colleagues. ^23^ We employed the potential outcomes framework for causal inference and made three standard assumptions, namely the ‘*no interference*’, ‘*no intervention anticipation*’ and ‘*no reversibility of intervention*’ assumptions. Under these assumptions, the potential outcomes framework posits that for every ODN *i* (*i* = 1, …, *n*) and month *t* (*t* = 1, …, *T*) there exists a set of potential outcomes 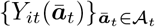, where ā_*t*_ = (*a*_1_, …, *a*_*t*_)^⊤^ is the intervention path i.e. a *t*-vector containing the intervention intensity (number of peers) at times 1, …, *t*. 𝒜_*t*_ is the set of all possible intervention paths up to time *t*, and *Y*_*it*_(ā_*t*_) is the outcome that we would observe in ODN *i* at time *t* if the peers intervention was unrolled as ā_*t*_. Let **0**_*t*_ = (0, …, 0)^⊤^ be the intervention path under which no peers are hired, that is ā_*t*_ = (0, …, 0)^⊤^.

For any ā_*t*_ ∈ 𝒜_*t*_, we let

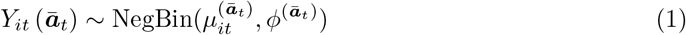

that is, the *negative binomial* distribution with mean 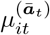 and variance 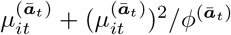.

We assumed that

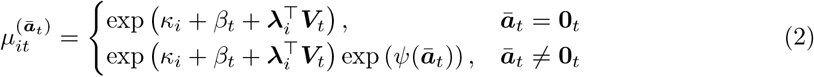

and

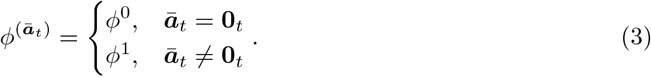

In Equation (2), *κ*_*i*_ are the ODN-specific intercepts that capture time-constant differences between ODNs, *β*_*t*_ are the time-fixed effects that capture temporal trends common to all ODNs, 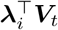 are interactive fixed effects allowing us to adjust for the time-varying effect of unobserved variables, and *ψ*(*ā*_*t*_) is the term that captures the effect of implementing the intervention according to intervention path ā_*t*_ on the outcome. We further assumed that *ψ*(·) is a smooth function of the exposure intensity (defined as cumulative number of peer-months) and its level depends on the presence of a lockdown. To achieve this, we let

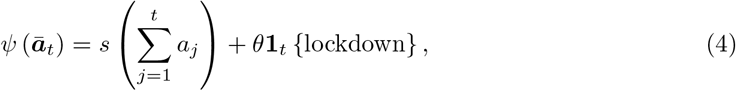

where *s*(·) is modelled using *B*-splines, and **1**_*t*_ lockdown equals one if the lockdown is in effect at time point *t* and zero otherwise. To complete the model specification for the potential outcomes, we assumed that potential outcomes under intervention 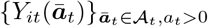 are independent of the intervention-free potential outcome *Y*_*it*_(**0**_*t*_) given model parameters and data.

Let *A*_*it*_ the observed number of peers in ODN *i* at time *t* and let *Ā*_*it*_ = (*A*_*i*1_, …, *A*_*it*_)^⊤^ be the intervention path that the ODN *i* has employed up to time *t*. We now specify the functional form for the assignment of the intervention path. For all *i* and *t*,

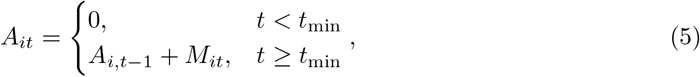

where *t*_min_ is the earliest time point at which a peer could be introduced, *M*_*it*_ is the total number of peers recruited by unit *i* at time *t*, and

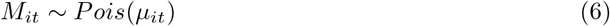

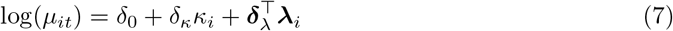

This assumption encodes our prior belief that in an observational study, assignment of the intervention may depend on a unit’s characteristics.

Let *Y*_*it*_ be observed outcome in ODN *i* at time *t*. The link between the potential outcomes and the observed outcomes is formalised through the consistency assumption, which states that

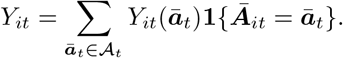

This allowed us to fit the model of Equations (1)-(7) to the data. We did this under the Bayesian paradigm using weakly informative priors, see full prior specifications in Schmidt and colleagues. ^23^ Samples from the posterior distribution of the model parameters were obtained using Markov chain Monte Carlo, specifically the No-U-Turn-Sampler as implemented in `R`package `rstan`. ^28^

We assessed the effectiveness of the peers intervention considering various *causal estimands* i.e. contrasts between potential outcomes (or their expectations). For any *t* and *i* such that *A*_*it*_ *>* 0, we defined the *individual intervention effect* (IIE) as

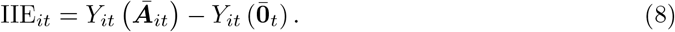

In words, IIE_*it*_ represents the number of additional HCV cases identified (or SVRs/drug service referrals) in ODN *i* at time *t* due to following intervention path *Ā*_*it*_ compared to the scenario in which the ODN did not introduce any peers up to time *t*. Let 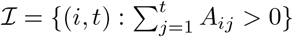 be the set of indices corresponding to ODNs that have experienced the peers intervention up to time *t*. The total number of additional HCV cases identified (or SVRs/drug service referrals) over the entire period due to the peers intervention is

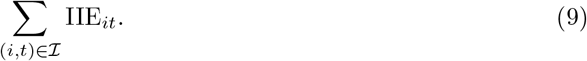

The total number of additional HCV cases identified during the lockdown period is defined analogously. The percentage increase in HCV cases identified (or SVRs/drug service referrals) thanks to the peers is

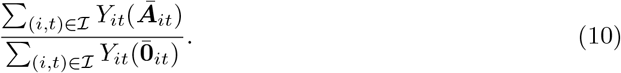

Finally, we considered the rate ratio of outcomes under intervention compared to no intervention given no lockdown is in place, defined as

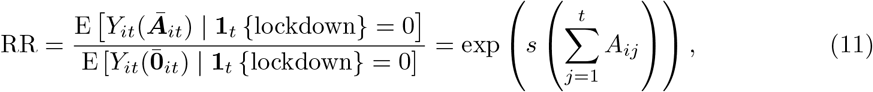

where the second equality follows from Equation (2). We note that the rate ratio is dependent on the cumulative exposure intensity; we therefore show it as a function of this quantity in the main text. The posterior distribution of all the estimands defined above can be obtained easily from the posterior distribution of the model parameters. For the RRs, it suffices to transform the posterior samples of *s*(*·*) as in Equation (11). For IIE, one needs to obtain draws from the posterior predictive of the counterfactuals 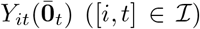. ^23^ Draws from the posterior predictive distribution of 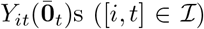 can be transformed into draws from the posterior distribution of IIE_*it*_s ([*i, t*] ∈ ℐ) using Equation (8). Draws from the posterior distribution of IIE_*it*_s can be aggregated into draws from the posterior distribution of the summary intervention effects using Equations (9) or (10).

